# Characteristics and outcomes of pregnant women hospitalised with confirmed SARS-CoV-2 infection in the UK: a national cohort study using the UK Obstetric Surveillance System (UKOSS)

**DOI:** 10.1101/2020.05.08.20089268

**Authors:** Marian Knight, Kathryn Bunch, Nicola Vousden, Eddie Morris, Nigel Simpson, Chris Gale, Patrick O’Brien, Maria Quigley, Peter Brocklehurst, Jennifer J Kurinczuk

## Abstract

**Objective:** To describe a national cohort of pregnant women hospitalised with SARS-CoV-2 infection in the UK, identify factors associated with infection and describe outcomes, including transmission of infection, for mother and infant.

**Design:** Prospective national population-based cohort study using the UK Obstetric Surveillance System (UKOSS).

**Setting:** All 194 obstetric units in the UK

**Participants:** 427 pregnant women admitted to hospital with confirmed Sars-CoV-2 infection between 01/03/2020 and 14/04/2020. 694 comparison women who gave birth between 01/11/2017 and 31/10/2018.

**Main outcome measures:** Incidence of maternal hospitalisation, infant infection. Rates of maternal death, level 3 critical care unit admission, preterm birth, stillbirth, early neonatal death, perinatal death; odds ratios for infected versus comparison women.

**Results:** Estimated incidence of hospitalisation with confirmed SARS-CoV-2 in pregnancy 4.9 per 1000 maternities (95%CI 4.5-5.4). The median gestation at symptom onset was 34 weeks (IQR 29-38). Black or other minority ethnicity (aOR 4.49, 95%CI 3.37-6.00), older maternal age (aOR 1.35, 95%CI 1.01-1.81 comparing women aged 35+ with those aged 30-34), overweight and obesity (aORs 1.91, 95%CI 1.37-2.68 and 2.20, 95%CI 1.56-3.10 respectively compared to women with a BMI<25kg/m^2^) and pre-existing comorbidities (aOR 1.52, 95%CI 1.12-2.06) were associated with admission with SARS-CoV-2 during pregnancy. 247 women (58%) gave birth or had a pregnancy loss; 180 (73%) gave birth at term. 40 (9%) hospitalised women required respiratory support. Twelve infants (5%) tested positive for SARS-CoV-2 RNA, six of these infants within the first 12 hours after birth.

**Conclusions:** The majority of pregnant women hospitalised with SARS-CoV-2 were in the late second or third trimester, supporting guidance for continued social distancing measures in later pregnancy. Most had good outcomes and transmission of SARS-CoV-2 to infants was uncommon. The strong association between admission with infection and black or minority ethnicity requires urgent investigation and explanation.

**Study Registration:** ISRCTN 40092247

## Introduction

The World Health Organation declared a global pandemic of coronavirus disease (SARS-CoV-2) in March 2020.^1^ As the number of confirmed cases increases, evidence on transmission, incidence and impact of SARS-CoV-2 infection in mothers and their babies remains limited. Pregnant women are not thought to be more susceptible to the infection than the general population.^2 3^ However, changes to the immune system mean that pregnant women may be more vulnerable to severe infection.^4^ Evidence from other similar viral illnesses such as influenza A/H1N1,^5–8^ SARS-CoV^9^ and MERS-CoV^10 11^ suggest that pregnant women are at greater risk of severe maternal and neonatal morbidity and mortality. There is some evidence that the risk of critical illness may be greatest in the later stages of pregnancy.^5 10 11^

To the best of our knowledge, as of April 16^th^ 2020 there had been more than 40 scientific reports of SARS-CoV-2 infection in pregnancy published in English, ^2 10 12 13^ none of which were population-based. The majority of reported cases occurred at term and women were delivered by caesarean section predominantly for maternal indication, although at least three studies reported cases of fetal distress.^13–15^ Most women developed mild or moderate symptoms including cough, fever and breathlessness, and only a small number developed severe disease.^15–19^. Risk factors were suggested to mirror those in the general population with one of the largest case series to date (n=43) reporting that nearly two thirds of cases had a BMI ≥ 30 kg/m^2^ and nearly half had comorbidities such as asthma (19%), Type 2 Diabetes (7%) or chronic hypertension (7%).^15^

The majority of neonates born to mothers with confirmed SARS-CoV-2 infection were asymptomatic and discharged home well. A small number of neonates had symptoms with a minority requiring admission to neonatal specialist care^14 15^, with only a few instances of reported transmission of SARS-CoV-2 infection to the neonate.^20–23^ Three neonates had elevated serum IgM antibodies identified shortly after birth in umbilical blood, but Sars-CoV-2 was not identified in any of these infants in the neonatal period despite testing. ^21 23^ These three infants were all asymptomatic, therefore the significance of vertical transmission remains unknown.

The aim of this study was to describe, on a population-basis, the risk factors, characteristics and outcomes of pregnant women hospitalised with SARS-CoV-2 in the UK, in order to inform ongoing guidance and management. This study was designed in 2012 and hibernated pending a pandemic, and was activated by the UK Department of Health and Social Care as an urgent public health study in response to the SARS-CoV-2 pandemic.

## Methods

A national prospective observational cohort study was conducted using the UK Obstetric Surveillance System (UKOSS).^24^ UKOSS is a research platform which collects national population-based information about specific severe pregnancy complications from all 194 hospitals in the UK with a consultant-led maternity unit. Nominated reporting clinicians were asked to notify all pregnant women with confirmed SARS-CoV-2 admitted to their hospital, using a live reporting link specific to each individual reporter. At the time covered by the study, that women were only tested if symptomatic for SARS-CoV-2 infection. The process was enabled by research midwives and nurses from the UK’s National Institute of Health Research Clinical Research Network following its adoption as an urgent public health priority study.^25^ In addition, nominated clinicians were sent a reporting email at the end of the month to ensure all cases had been reported, and to confirm zero reports (active negative surveillance). Following notification, clinicians were asked to complete an electronic data collection form containing details of each woman’s characteristics, management and outcomes. Reporters who had not returned data were contacted by email at week one, two and three after notification. This analysis reports characteristics and outcomes of women who were notified as hospitalised 01 March- 14 April 2020 and for whom complete data had been received by 29 April 2020.

Patients and the public were involved in the design of the study, and, as part of the UKOSS Steering Committee, in the conduct of the study and interpretation of the results.

Information about a comparison cohort of women was obtained from a previous study of seasonal influenza in pregnancy.^26^ Comparison cases were the two women giving birth immediately prior to any woman hospitalised with confirmed influenza between 01 November 2017 and 30 October 2018. A historical comparison cohort was used to ensure there was no possibility that comparison women had asymptomatic or minimally symptomatic SARS-CoV-2 infection. Data on maternal and perinatal deaths were cross-checked with data from the MBRRACE-UK collaboration, the organisation responsible for maternal and perinatal death surveillance in the UK. ^27^

### Sample size

In this national observational study, the study sample size was governed by the disease incidence, thus no formal power calculation was carried out. With 427 women in the exposed (hospitalised with SARS-CoV-2 infection) cohort and 694 women in the comparison cohort, this analysis had 80% power at p<0.05 to detect a 1.5 fold or greater difference in the prevalence of a more common risk factor (25% prevalence amongst comparison women) as statistically significantly different, and a 2.0 fold or greater difference in a less common risk factor (5% prevalence).

### Statistical analysis

The incidence of hospitalisation with confirmed SARS-CoV-2 infection in pregnancy was calculated using denominator estimates based on the most recently available (2018) national maternity data for the constituent countries of the United Kingdom. Numbers and proportions are presented with 95% confidence intervals. Women hospitalised with Sars-CoV-2 were compared with women in the comparison cohort using univariable and multivariable unconditional logistic regression to estimate odds ratios with 95% confidence intervals (CI). A sensitivity analysis was undertaken excluding women from London, West Midlands and the North West of England to explore whether factors associated with admission with infection varied outside of the populations with the highest general rates of infection. Variables that were significantly different between groups were included as confounders in the analyses comparing maternal and perinatal outcomes between groups. Statistical tabulation and analyses were performed using STATA version 15 (KB/MK).

### Study registration

The study is registered with ISRCTN, number 40092247, and is still open to case notification. The study protocol is available at https://www.npeu.ox.ac.uk/ukoss/current-surveillance/covid-19-in-pregnancy.

## Results

Responses were received from all 194 hospitals with obstetric units in the UK (100%). Through 1 March to 14 April 2020, 630 women hospitalised with confirmed SARS-CoV-2 infection in pregnancy were notified in the UK, among an estimated 86,293 maternities. Data were returned for 579 women (92%); 15 were duplicate cases, 35 reported in error, 87 were diagnosed as outpatients and not admitted overnight, 9 had no positive PCR test and no evidence of pneumonitis on imaging, and 6 had no evidence of infection during pregnancy, leaving 427 pregnant women hospitalised with confirmed SARS-CoV-2 across the UK. This represents an estimated incidence of 4.9 per 1000 maternities (95%CI 4.5-5.4 per 1000 maternities).

Women were symptomatic at a median of 34 completed weeks’ gestation (IQR 29-38), with the majority of hospitalised women symptomatic in the third trimester of pregnancy or peripartum (n=342/424, 81%). The commonest symptoms reported by women were fever, cough and breathlessness (Figure 1). Black and other minority ethnicity, the presence of pre-existing comorbidity, older maternal age and overweight or obesity were all associated with admission with SARS-CoV-2 infection in pregnancy (Table 1). The association with black or other minority ethnicity persisted in the sensitivity analysis (OR 3.67, 95%CI 2.55-5.28).

**Figure 1:**
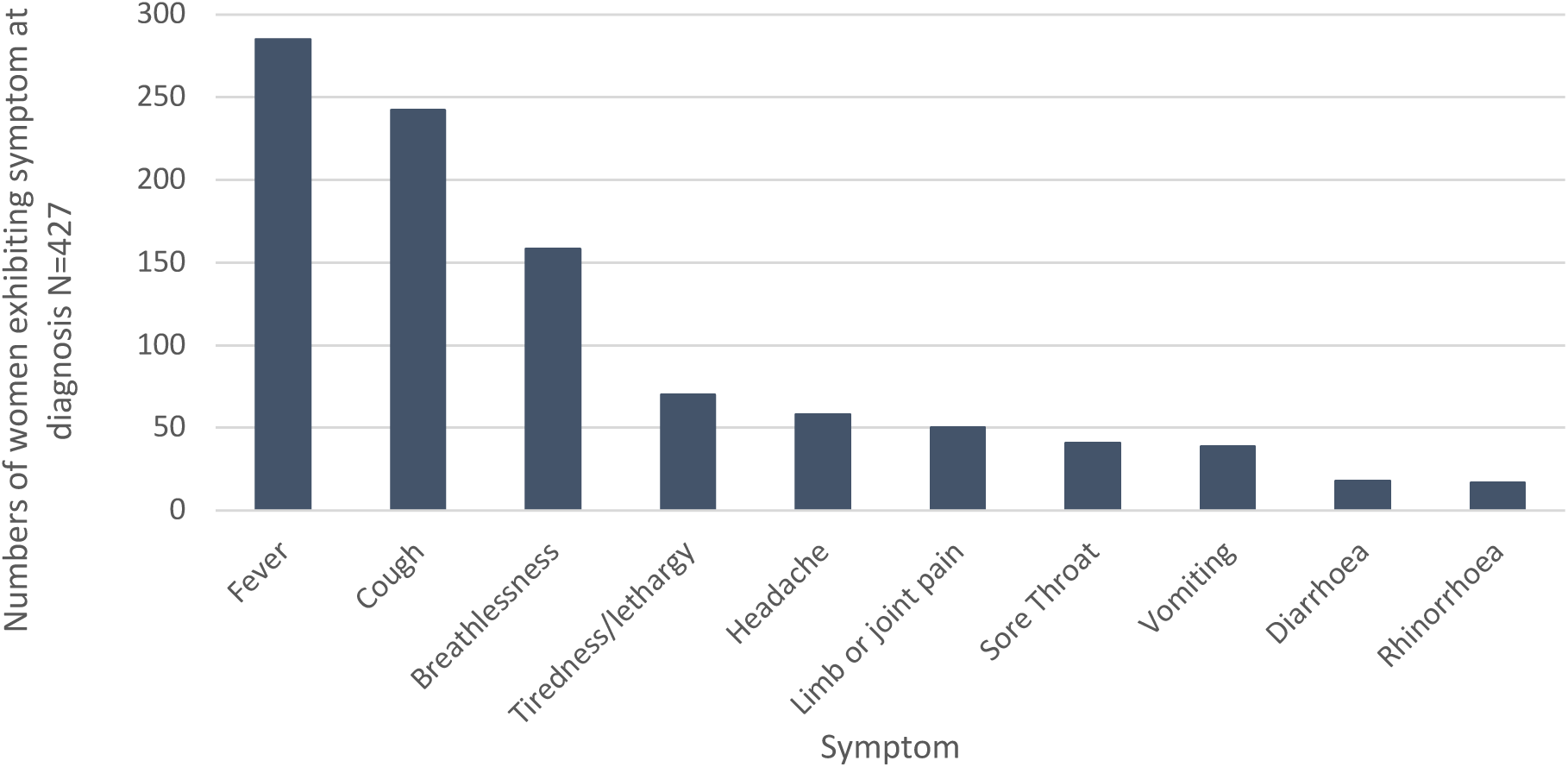
Maternal symptoms at diagnosis.

**Table 1:**
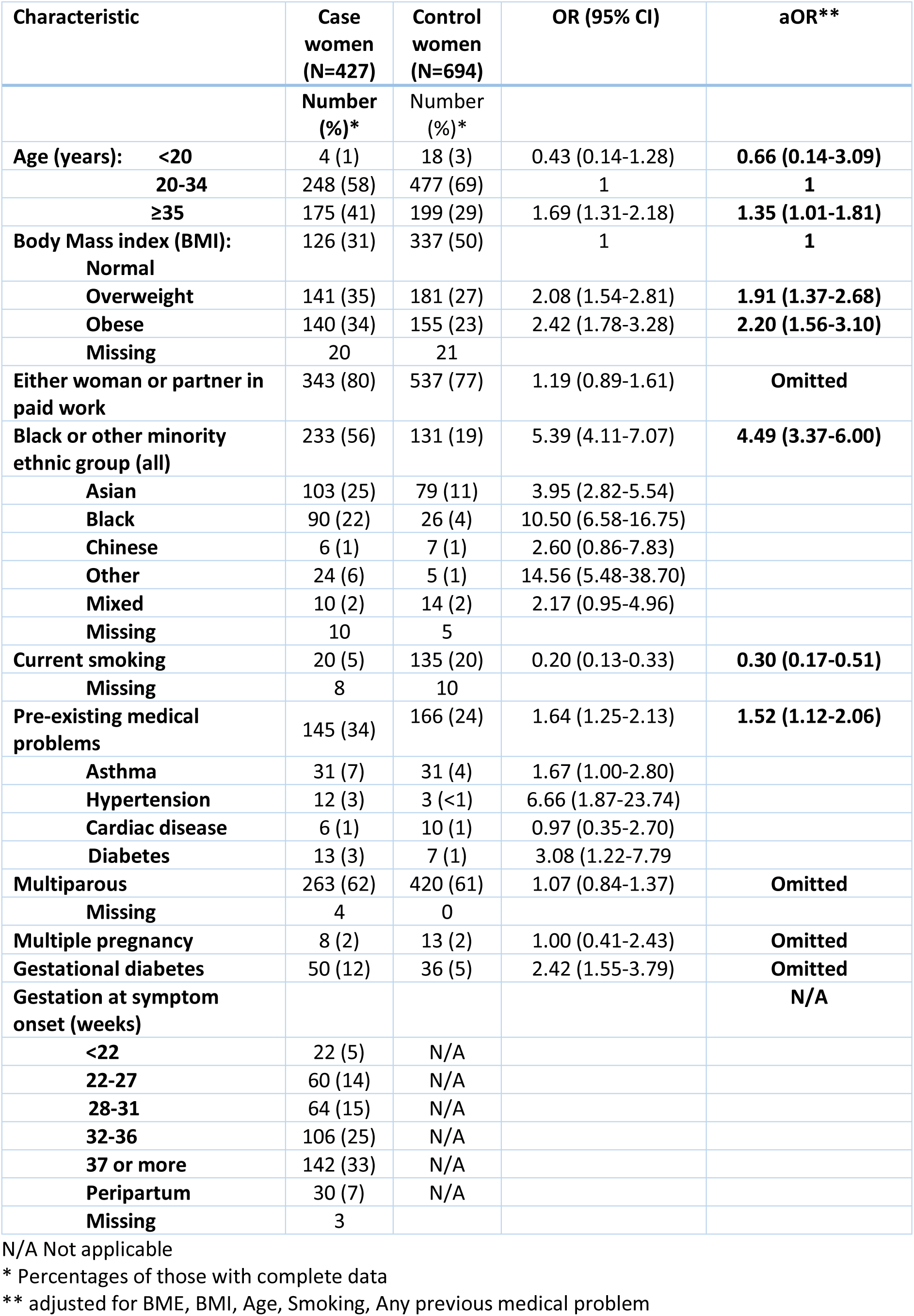
Characteristics of pregnant women with confirmed SARS-CoV-2 infection for whom data were available, UK, 01 March to 14 April 2020 and control women.

Two hundred and forty-seven women (58%) hospitalised gave birth or had a pregnancy loss; the remaining 180 (42%) women had ongoing pregnancies at the time of this analysis. Forty (9%) women required level 3 critical care; four of these women received extracorporeal membrane oxygenation (Table 2). Of the women who received critical care, 31 had been delivered due to COVID-19, nine were still pregnant. Eight (89%) of the women who were pregnant at the time of their critical care admission had been discharged. Fifteen (48%) of the postnatal women had been discharged at the time of this analysis, three had died and 13 were still inpatients, of whom nine remained in critical care. Overall, five women who were admitted and had a positive test for SARS-CoV-2 died, a case fatality of 1.2% (95% CI 0.4-2.7%) and a SARS-CoV-2-associated maternal mortality rate of 5.6 (95% CI 1.8-13.1) per 100,000 maternities. In total, thirty women (7%), 10 antenatal and 20 postnatal, were still inpatients at the time of this analysis.

**Table 2:** Hospital outcomes and diagnoses amongst women with confirmed SARS-CoV-2 infection in pregnancy.

| **Maternal outcomes** | **Case women (N=427)** | **Comparison women (N=694)** | **OR (95% CI)** | **aOR (95% CI)\*** |
| --- | --- | --- | --- | --- |
|  | **Number (%)** | **Number (%)** |  |  |
| **Required critical care** | 40 (9) | 1 (<1) | 76.89 (10.53-561.6) | 58.61 (7.81-439.8) |
| **Required ECMO** | 4 (1) | 0 |  |  |
| **SARS-CoV-2 pneumonia on imaging** | 101 (24) | - |  |  |
| **Final outcome** |  |  |  |  |
| **Died** | 5 (1) | 0 (-) |  |  |
| **Discharged well** | 392 (92) | n/a |  |  |
| **Still admitted** | 30 (7) | n/a |  |  |

\*Adjusted for maternal age, BMI, ethnicity and presence of co-morbidities

Nine women (2%) were treated with an antiviral, eight of whom were given oseltamivir, one of whom also received lopinavir/ritonavir. One woman was given remdesivir. Sixty-one women (14%) were given corticosteroids for fetal lung maturation, of whom 40 (66%) went on to give birth.

Of the 243 women who had given birth, 180 (74%) gave birth at term (Table 3). Sixty-three women gave birth preterm; 50 had iatrogenic preterm births (79%), 29 (46%) due to maternal COVID-19, nine (14%) due to fetal compromise and twelve (19%) due to other obstetric conditions. Fifty-nine percent of women (n=144) had a caesarean birth, but the majority of caesarean births occurred for indications other than maternal compromise due to SARS-CoV-2 infection. Thirty-nine women (27% of those having a caesarean birth) had a caesarean birth for reasons of maternal compromise, 34 (24%) for concerns about fetal compromise, 28 (19%) due to failure to progress in labour or failed induction of labour, 21 (15%) for other obstetric reasons, 16 (11%) because of previous caesarean birth and 6 (4%) at maternal request. Twenty-eight women (20%) had general anaesthesia for their caesarean birth; 18 (64%) of these women were intubated due to maternal respiratory compromise and ten (36%) were intubated to allow for urgent delivery. Twenty-nine percent of comparison women (n=200) had a caesarean birth, 13 of whom (7%) had general anaesthesia.

**Table 3:** Pregnancy and infant outcomes amongst pregnant women with confirmed SARS-CoV-2 infection.

| Pregnancy outcomes | Case women<br>(N=427) | Comparison<br>women<br>(N=694) | OR (95% CI) | aOR (95%<br>CI) |
| --- | --- | --- | --- | --- |
|  | **Number (%)** | **Number (%)** |  |  |
| Ongoing pregnancy | 180 (42) |  |  |  |
| Pregnancy completed | 247 (58) | 694 |  |  |
| Pregnancy loss | 4 (1) | 2 (<1) | 3.27 (0.60-17.94) | 3.79 (0.46-30.94) |
| Stillbirth | 3 (1) | 2 (<1) | 2.46 (0.41-14.81) | 3.48 (0.54-22.26) |
| Live birth | 240 (97) | 690 (99) | 0.23 (0.04-1.40) | 0.16 (0.02-1.01) |
| Neonatal death | 2 (1) | 1 (<1) | 5.73 (0.52-63.52) | 4.24 (0.34-53.57) |
| Gestation at end of pregnancy<br>(weeks) |  |  |  |  |
| <22 | 4 (2) | 2 (<1) |  |  |
| 22-27 | 5 (2) | 5 (1) |  |  |
| 28-31 | 17 (7) | 6 (1) |  |  |
| 32-36 | 41 (17) | 44 (6) |  |  |
| 37 or more | 180 (73) | 636 (92) |  |  |
| Median (IQR) | 38 (36-40) | 39 (38-40) |  |  |
| Mode of birth |  |  |  |  |
| Caesarean – maternal<br>indication due to SARS-CoV-2 | 39 (16) | 0 (0) |  |  |
| Caesarean – other<br>indication | 105 (43) | 201 (29) |  |  |
| Operative vaginal | 25 (10) | 74 (11) |  |  |
| Unassisted vaginal | 76 (31) | 417 (60) |  |  |

\*Adjusted for maternal age, BMI, ethnicity and presence of co-morbidities

**Table 4:** Infant outcomes amongst live born babies of women with confirmed SARS-CoV-2 infection in pregnancy.

| Infant outcomes | Live born infants of<br>women with Sars-CoV-2 (N=244) | Infants of<br>comparison women<br>(N=703) |
| --- | --- | --- |
|  | **Number (%)** | **Number (%)** |
| NICU Admission | 64 (26) | 37 (5) |
| Positive SARS-CoV-2 test (Liveborn infants only) |  |  |
| No | 232 (95) | N/A |
| Positive test <12 hrs of age | 6 (2) | N/A |
| Positive test ≥12 hrs of age | 6 (2) | N/A |

Five babies died; three were stillborn and two died in the neonatal period. Three deaths were definitely unrelated to SARS-CoV-2 infection, for two stillbirths it was unclear whether SARS-CoV-2 contributed to the death. Sixty-four infants (26%) were admitted to a neonatal intensive care unit (NICU), 46 of whom (72%) were preterm, including 19 (29%) who were<32 weeks’ gestation. One infant was diagnosed with neonatal encephalopathy (grade 1) after a spontaneous vaginal birth at term. Twelve (5%) infants of women hospitalised with infection tested positive for SARS-CoV-2 RNA, six of these infants within the first 12 hours after birth. Two of the six infants with early onset SARS-CoV-2 infection were unassisted vaginal births, four were born by caesarean, three of which were pre-labour. The six infants who developed later infection were born by pre-labour caesarean (n=4) and vaginal birth (n=2). Only one of the infants with an early positive test for SARS-CoV-2 RNA was admitted to a neonatal unit, compared to five infants with a later positive test.

## Discussion

The clinical data from this national surveillance study show black and minority ethnicity is significantly associated with admission with SARS-CoV-2 infection in pregnancy, along with preexisting co-morbidities, overweight and obesity and older maternal age. Women admitted with infection were less likely to smoke. About one in 10 pregnant women admitted to hospital in the UK with SARS-CoV-2 infection required respiratory support in a critical care setting. These patterns are similar to those in the general population hospitalised with Sars-CoV-2. ^28^ Over half of all women admitted with SARS-CoV-2 infection have given birth; 12% were delivered preterm solely due to maternal respiratory compromise. Almost sixty percent of women gave birth by caesarean section; the majority of caesarean births were for indications other than maternal compromise due to SARS-CoV-2 infection. One in twenty of the babies of mothers admitted to hospital subsequently had a positive test for SARS-CoV-2; half had infection diagnosed on samples taken at less than 12 hours after birth.

This rapid report has been produced at a time when active transmission of SARS-CoV-2 is still occurring, with around 100 pregnant women hospitalised in the UK with infection each week, and the limitations of these data must therefore be recognised. We do not yet have complete pregnancy outcomes for women who were admitted but subsequently discharged well, and a number of women were still inpatients at the time of writing. The data collected for this rapid national cohort study were restricted to essential items, therefore we do not have daily indicators of women’s clinical condition, blood and other test results. Nevertheless, these results do show the benefits of systems such as UKOSS which can be rapidly activated to undertake comprehensive studies such as this in a public health emergency. UKOSS studies were activated for influenza A/H1N1 ^33^ and Zika virus^34^ in pregnancy; countries in the International Network of Obstetric Survey Systems (INOSS) ^35^ are also undertaking similar national studies to allow for the unification of population-based data across multiple countries and avoiding the biases of data collected through centre-based registries. The National Institute for Health Research’s Clinical Research Network, ^36^ with midwifery and obstetric leads coordinating networks of research staff, also help ensure rapid and accurate collection of these valuable data even in the context of the pressurised health system in a pandemic.

The addition of these national, population-based data to existing reports provides clarity on the outcomes of infection in pregnant women. Previous published information has been largely based on individual hospital case series, cases identified across small series of hospitals but with a lack of clarity on the proportion of cases ascertained, or from registry data which are also likely to contain information on a non-representative series of cases, with problems of overlap and duplicate reporting; population-based data are essential to provide unbiased information on incidence and outcomes. During the period these data were collected around 90,000 women gave birth in the UK; 427 were notified as having been admitted with SARS-CoV-2 in pregnancy; fewer than 1 woman hospitalised for every 200 women giving birth. Approximately one woman per 2400 giving birth required critical care admission. The overall maternal mortality rate with confirmed SARS-CoV-2 infection was around 1 in 18,000 women giving birth.

The association between black and minority ethnicity and hospitalisation with SARS-CoV-2 in pregnancy is of concern and requires further investigation. Our sensitivity analysis suggests that this cannot simply be explained by higher incidence in the main metropolitan areas with higher proportions of women from ethnic minority groups, as evidence of effect persisted when women from London, the West Midlands and the North West were excluded. The effect also persisted despite adjustment for age, BMI and co-morbidities. Ethnic disparities in incidence and outcomes have been noted amongst non-pregnant populations with SARS-CoV-2 infection, notably in the US ^29^, and a range of possible reasons have been suggested for these observed disparities, including social behaviours, health behaviours, co-morbidities and potentially genetic influences ^30^. Health system factors have been suggested to underlie the disparity in the US; the fact that these disparities exist in a country with a universal free to access health care system indicate that the health system cannot be the sole explanation.

In common with previous reports, the majority of women hospitalised with SARS-CoV-2 in pregnancy were in the late second or third trimester, which replicates the pattern seen in other respiratory viruses of women in later pregnancy being more severely affected. This supports the current guidance for strict social distancing measures amongst pregnant women, particularly in their third trimester ^2^. It should be noted, however, that higher hospitalisation rates in the third trimester were also reported in the context of influenza,^31^ and thought to be for precautionary reasons, rather than necessarily because of maternal compromise. Although case notification has been augmented through a link with the UK Early Pregnancy Surveillance System (UKEPSS), ^32^ it is also possible that the route of identification of the cases included in this series, through UK obstetric units, has led to under-ascertainment of women admitted in the early stages of pregnancy.

Outcomes for infants are largely reassuring when considering potential impacts of SARS-CoV-2 infection acquired before or during birth; the small number of early PCR positive infants of mothers with infection did not have evidence of severe illness. This observation of only mild disease has also been reflected in early case reports of infant infection in the perinatal period. ^20–23^ Nevertheless, 2% of infants did have evidence of viral RNA on a sample taken within 12 hours of birth, which suggests that vertical transmission may be occurring. We have no evidence as to whether IgM was raised in these infants and therefore whether infection was acquired before or during birth, but three infants tested positive following pre-labour caesarean section. During the study period UK guidance for postnatal management of infants born to mothers with confirmed or suspected SARS-CoV-2 infection was to keep mother and infant together and to encourage breastfeeding with consideration of using a fluid-resistant surgical face mask for the mother. These findings emphasise the importance of infection control measures around the time of birth, and support the advice given by the World Health Organisation around precautions to take whilst breastfeeding.

We conducted this study in a high resource setting with universal healthcare free at the point of access, and findings would therefore be generalisable to similar settings. The fact that the majority of women experience mild infection would suggest that outcomes are likely to be good in settings with less well developed health systems. However, given the proportion of women admitted who required critical care, it is likely that the outcomes of severe infection will be poorer in the absence of such facilities.

In the context of the Sars-CoV-2 pandemic, ongoing data collection about the outcomes of infection during pregnancy will remain important. There remain unanswered questions about the extent and impact of asymptomatic or mild infection. Serological studies, as well as those using retrospective data to identify women with either confirmed or presumed mild infection in pregnancy, will be essential to fully assess potential impacts such as congenital anomalies, miscarriage or intrauterine fetal growth restriction. Nevertheless, these data suggest that the majority of women do not have severe illness and that transmission of infection to infants of infected mothers may occur but is uncommon.

## Data Availability

Data will be shared according to the National Perinatal Epidemiology Unit data sharing policy https://www.npeu.ox.ac.uk/downloads/files/npeu/policies/Data%20Sharing%20Policy.pdf

## Author contributions

MK wrote the first draft of the article with contributions from NV and KB. KB and MK carried out the analyses. All authors edited and approved the final version of the article. MK, NS, EM, JJK, CG, MQ, PO’B, PB, KB contributed to the development and conduct of the study.

## Acknowledgements

The authors would like to acknowledge the assistance of UKOSS and UKEPSS reporting clinicians, the UKOSS Steering Committee, The NIHR Clinical Research Networks and UKEPSS leads at the University of Birmingham, without whose support this research would not have been possible. We would like to acknowledge the help and support of the UKOSS admin team: Melanie O’Connor and Anna Balchan, and additional staff who assisted on a temporary basis: Madeleine Hurd, Victoria Stalker, Alison Stockford and Rachel Williams.

## Ethics committee approval

This study was approved by the HRA NRES Committee East Midlands – Nottingham 1 (Ref. Number: 12/EM/0365).

## Data Sharing Statement

Data from this study will be shared according to the National Perinatal Epidemiology Unit Data Sharing Policy available at: https://www.npeu.ox.ac.uk/downloads/files/npeu/policies/Data%20Sharing%20Policy.pdf

